# Cohort profile: design and methods for Project HERCULES (Healthcare Exemplar for Recovery from COVID 19 by Use of Linear Examination Systems). Multi-disciplinary implementation and evaluation of an asynchronous review clinic for monitoring chronic eye disease in English NHS services

**DOI:** 10.1101/2024.11.05.24316762

**Authors:** Dhakshi Muhundhakumar, Caroline S Clarke, Grant Mills, Angus I G Ramsay, Kerstin Sailer, Peter Scully, Duncan Wilson, Dun Jack Fu, Siyabonga Ndwandwe, Rosica Pachilova, Anne Symons, Steve Napier, Joy Adesanya, Gus Gazzard, Robin Hamilton, Jonathan Wilson, Paul Webster, Peng T. Khaw, Sobha Sivaprasad, Hari Jayaram, Paul J. Foster, HERCULES Consortium, HERCULES Consortium

**Author notes:** Joint senior authors, taking joint credit and responsibility (Sobha Sivaprasad, Hari Jayaram, Paul J. Foster). Correspondence to: Professor Paul Foster.

## Abstract

**Purpose:** To describe the research principles and cohort characteristics of the multidisciplinary Project HERCULES, which evaluated implementation of an innovative model of high-volume outpatient eyecare service to monitor patients with stable chronic eye diseases. The rationale was to improve capacity and efficiency of eyecare in the National Health Service (NHS) in England through the creation of technician-delivered monitoring in a large retail-unit in a London shopping-centre, with remote asynchronous review of results by clinicians (named Eye-Testing and Review through Asynchronous Clinics (Eye-TRACs)). UCL’s Bartlett School of Sustainable Construction produced the RIBA (Royal Institute of British Architects) Stage 1 briefing requirements for optimal design specifications and operational parameters for this new model of care from first principles research, by analysing and developing ergonomic data from multiple iterations.

**Participants:** Patients aged 18 years or above being monitored in secondary care in Moorfields Eye Hospital NHS Trust for stable glaucoma or retinal conditions were given appointments at Eye-TRAC at Brent Cross, London. Willing participants were recruited when attending Eye-TRAC from September 2021-November 2023 and formed the ‘intervention cohort’. The ‘comparator cohort’ consisted of patients that continued to be monitored in secondary care during the same period. Other than residence of the participants, there were no other demographic or disease severity differences in this cohort. Additionally, anonymised data from across the Trust informed an analysis of the impact of opening the Eye-TRACs on Trust-wide waiting times. A nationwide stakeholder preference survey of health-care professionals, members of the public and ophthalmology patients was carried out.

**Findings to date and conclusion:** 41,567 patients attended the Brent Cross Eye-TRAC between September 2021 and November 2023. 5,539 patients were recruited to Project HERCULES. Four spatial “iterations,” with different configurations of equipment were investigated in succession. Spatial configurations promoting independently parallel patient journeys with limited queuing, and direct line of sight between diagnostic stations, supported efficient patient flow. The latter iteration incorporated cataract clinics. Although it added more system complexity, it enabled the evaluation of a further indication for use of Eye-TRAC.

**Future plans:** Qualitative analysis of patient and staff feedback alongside rapid ethnographic work to streamline services is under way. We seek to develop a framework to help inform NHS guidance for ophthalmology and other outpatient diagnostic services. Our data will be analysed to identify enhancements to further streamline operational efficiency. We will identify and enumerate limitations in information technology that create bottle-necks in the review process.

**Sponsor:** Moorfields Eye Hospital NHS Foundation Trust

**Sponsor protocol reference:** JAYH1011

**Integrated Research Application System (IRAS) ID:** 303760

**Funders:** NIHR Biomedical Research Centre at Moorfields Eye Hospital NHS Foundation Trust & UCL Institute of Ophthalmology, London, UK

Moorfields Eye Hospital NHS Foundation Trust Moorfields Eye Charity

Ubisense Ltd, Zeiss, Optos

**STRENGTHS AND LIMITATIONS of this study:**

- Our multi-disciplinary research team is a major strength of the work; new collaborations and understandings have arisen that cut across academic disciplines and we hope this will provide meaningful lessons for health services now and in future.
- Design development that used rapid experimentation to test new ideas (before spending significant resources on them) was employed; we collected data to build an evidence base, dynamically test new environments, build protypes and execute analysis iteratively.
- Continuous input from technicians, administrative and managerial staff led to improvements in later iterations and greater buy-in from staff and ultimately the success of the project.
- Patient and public involvement was integral to the design and development of the quantitative and qualitative work.
- Due to the need for rapid service capacity expansion and high-volume throughput (as a result of the pandemic) the conditions and comparisons within the study could not be tightly controlled.

## INTRODUCTION

### Impact of COVID-19 on outpatient ophthalmology services

The COVID-19 pandemic precipitated the greatest health crisis in living memory(1). The suspension of non-urgent NHS work led to millions more delayed appointments in an already strained eyecare service across the United Kingdom and, after years of health funding austerity, this backlog was potentially insurmountable. Overall, NHS England surgical waiting lists rose from 4.4 million pre-pandemic to over 8 million (December 2023) and have been projected by the National Audit Office to rise to 12 million by 2025(2). Ophthalmology, as the busiest outpatient specialty (with over 8 million annual attendances in the NHS in England (3)) was severely affected. Before the pandemic (2017), 3,384 ophthalmology patients in the UK suffered follow-up delays greater than a year, with 1% of these suffering preventable loss of vision(4). In 2020, the Health Services Safety Investigations Body published a report on lack of timely glaucoma monitoring and inadequate hospital eye service capacity after an incident where a 34-year-old woman became blind, and highlighted the need for hospital eye services to work innovatively to deliver timely care(5). Following the first lockdowns of the pandemic in March 2020, over one million NHS ophthalmology appointments were deferred, adding to the huge existing backlog and the potential for thousands of cases of avoidable blindness(5).

In 2013, direct costs of NHS eye care were £3 billion in the UK, with a further £6 billion/year of indirect costs of sight loss(6). People fear blindness more than severe angina or kidney dialysis(7). Patients suffering from chronic eye disease comprised a high-risk demographic for COVID-19 morbidity and mortality, most being elderly, and many with multiple systemic co-morbidities and disproportionately of minority ethnic heritage(8, 9).

### Rationale for a novel approach to monitoring chronic eye diseases

Glaucoma, age-related macular degeneration (AMD), diabetic retinopathy (DR) and cataract are increasing in prevalence due to an ageing population, and are being identified and referred earlier due to the availability of advanced imaging technologies such as optical coherence tomography (OCT) in high street optometry practices(10). These patients need regular imaging and functional testing to monitor their chronic and often lifelong eye conditions(11).

Under current care pathways, patients with stable glaucoma, AMD and DR attend outpatient appointments in secondary care. These appointments can take several hours, with large numbers of patients sitting in waiting rooms that are frequently cramped and poorly ventilated. During the appointment, they may interact with nurses, technicians, optometrists and doctors, with a waiting time between seeing each health professional and the various tests required. In view of the increasing capacity constraints, the dangers of COVID-19 and other air-borne disease transmission, and the vulnerable population that attend these clinics, it has become increasingly accepted that the traditional outpatient model of delivering care for chronic eye disease needs to be re-appraised(12, 13).

During the inception of Project HERCULES in April 2020, minimising the potential for COVID-19 transmission was the overriding priority. Since then, the focus has shifted to building more high-volume chronic eye disease monitoring capacity in the form of diagnostic and monitoring clinics to overcome backlogs and increase patient throughput within NHS services (14) including, more recently, cataract surgery.

In the context of significant capacity challenges, more efficient, higher volume services are needed to monitor chronic stable patients and identify those in need of treatment escalation(15). An adjunct to increased capacity is risk stratification, based upon relevant data, allowing the identification of higher risk patients who are more susceptible to harm from delayed appointments(16).

Asynchronous review, or “virtual” clinics (as they have been previously broadly referred to) are well established as a model of care(17–27) whereby patients undergo testing by technicians with results being reviewed by clinicians at a later time. Moorfields established “virtual diagnostic and monitoring clinics” in the main hospital in 2014 (Clinic 1A)(28), and thereafter expanded the service to other sites. They have become part of the “new normal” landscape of hospital ophthalmology services(13, 17–19, 29, 30), providing a route to increasing clinic capacity in the face of otherwise insurmountable backlogs(21, 22, 31, 32) for patients whose clinical condition is deemed “probably stable”(28). These clinics are led by ophthalmic technicians, with operational protocols designed to examine patients in a fraction of the time of a conventional hospital appointment, thus reducing staffing costs and increasing productivity. Test results are reviewed asynchronously and remotely by specialist clinicians(29, 33) enabling them to assess a larger number of patients per session when compared to the conventional face-to-face model. We refer to these as Eye-Testing and Review through Asynchronous Clinics (Eye-TRACs) in this paper.

### AIMS & OBJECTIVES

This project aimed to provide research evidence on how to build additional clinic capacity focussing on patients requiring monitoring for chronic eye diseases such as AMD, DR and glaucoma, and latterly added cataract services.

Within this laboratory of clinical efficiency, we gathered data on:

- Patient and staff movement through an examination pathway, using a combination of direct observation, real-time positional sensor tracking, equipment time stamps and administrative records of arrival and departure times, to identify and overcome capacity bottlenecks.
- How reconfiguration of the physical layout of the clinic influenced efficiency, performance and patient experience.
- The operational impact of introducing pre- and post-operative cataract services to the Eye-TRACs.

In parallel, we are reviewing the features associated with greater efficiency in existing high-volume cataract services in the UK and globally.

Through analysis of quantitative measures and qualitative feedback, we aimed to formulate an evidence-based toolkit to reproduce high-efficiency and high-satisfaction monitoring pathways for chronic conditions within global healthcare systems.

### COHORT DESCRIPTION

#### Settings

##### Project HERCULES Clinic – A Reconfigurable Clinical Innovation Laboratory

A retail unit in a major north London shopping mall (Brent Cross) was identified as a suitable site for an Eye-TRAC. Building on prior research and innovation (17–19, 21–27, 31), the Eye-TRAC was built in this rapidly converted retail space.

This new high throughput Eye-TRACs commenced with a linear flow design, followed by three further rapid iterations of physical layout and processes enabled by a reconfigurable partition system developed by the Bartlett School of Architecture which used Design for Manufacture principles (minimize the number of parts, use standardised parts, create modular designs, use sustainable components with short, wide supply chains) to incorporate innovations, such as clustered or zonal use of diagnostic equipment. We intended also to examine how we might improve efficiency through optimisation of the physical layout of the clinic while minimising the risk of aerosol transmission to vulnerable patients(34). The study commenced in October 2021 and closed recruitment in December 2023.

##### Alternative Comparator Clinics

Moorfields Eye-TRACs at Hoxton, Cayton Street and City Road. These are pre-existing asynchronous review clinics for staffle glaucoma and retina patients – they were previously known as “virtual clinics” but for consistency of terminology will be referred to as Eye-TRACs. The City Road Eye-TRAC was previously referred to as Clinic 1A.

##### Face to Face clinics

Traditional hospital clinic outpatient care at Moorfields City Road site. This was the commonest consultation type (~80%) before the pandemic.

#### Participant recruitment and assessment

This project was approved by the North East – York NHS Research Ethics Committee. Eligible patients were those attending glaucoma and retina Eye-TRACs or latterly cataract clinics over the age of 18 and living locally. Prior to arrival at the clinic, all eligible patients were sent information on our research. They were advised that they may be asked if they wished to participate in the research project, which would involve wearing a tracking device during their visit and completing patient-reported outcome measure (PROM) questionnaires including demographic, visual status and quality of life (QOL) information and other patient reported outcome measures (see appendix) at the end of their appointment, and they were free to decline with no alteration in the care they would receive. On arrival, eligible patients were offered the opportunity to participate in the research. Not all eligible patients were approached, as it depended on the availability of members of the research team.

All patients then passed through the clinic, starting their journey with a health status review by an ophthalmic technician. This was followed by assessments of visual acuity, visual fields, intraocular pressure, OCT imaging and widefield fundus photography. Cataract and retina patients underwent pharmacological pupil dilation following initial visual acuity and intraocular pressure assessment. The diagnostic equipment included Humphrey Field Analysers 3 (HFA3) and optical coherence tomography (OCT) devices (Cirrus OCT 6000, Zeiss, Germany, for glaucoma; Spectralis OCT, Heidelberg Engineering, Germany, for retina; and ultra-widefield retinal imaging devices from Optos, Dunfermline, UK).

The study focussed on the <u>following priority areas</u> within the Eye-TRACs:

##### Monitoring of Patient and Staff Movement

We aimed to capture directly-observed, real-time measures of test duration and overall patient journey across existing similar Moorfields clinics, and four different experimental iterations of the Eye-TRACs, in a subset of participants at each site. This was performed by participant observers from the UCL Bartlett School of Architecture. Additionally, at the HERCULES Eye-TRAC patient and staff movements within the clinical facility were tracked using ultra-wideband real-time location systems (Ubisense, Cambridge, UK) during the four iterative configurations of the diagnostic clinic. Patients and staff gave informed consent for this monitoring. Patients also gave consent to access to their basic relevant clinical data and were asked for consent to be contacted to provide feedback on patient panels and participate in future related projects.

##### Implementation, Layout Efficiency, Service Performance, Patient and Staff Experience

Researchers from the UCL Research Department of Behavioural Science and Health used qualitative approaches (stakeholder and non-participant interviews) to study implementation of the new service and how staff and patients experienced it. They carried out a rapid ethnographic study to explore adverse experiences of the service, such as excessively long appointments and missed diagnostic tests. They carried out semi-structured interviews with staff, attended briefing meetings before shifts and observed (with consent) staff-patient interactions, to develop guidance on how to avoid unintended outcomes.

The UCL Bartlett School of Sustainable Construction and UCL Bartlett Centre for Advanced Spatial Analysis investigated the features of the various clinic layouts that worked well and those that did not. This involved generating testaffle clinic design ideas, pre-testing the evidence through simulation, building prototypes, analysing the results using the data generated by the ultra-wideband real-time location system and sharing learning for redesigning. This work demonstrated the principles of building physical / digital twin clinics which allow the development of a “living lab” that can set the standard for a novel experimental approach to optimising layout and building designs for healthcare.

##### Quantitative Analyses of the Healthcare System Beyond Brent Cross

Health economists from the UCL Research Department of Primary Care and Population Health are using anonymised system-wide Moorfields patient-level data to evaluate the change in appointment delays (defined using the “latest clinically appropriate date” associated with each appointment) across Moorfields network sites over time, incorporating appointment information recorded in Moorfields routine electronic health records from 2018 to 2023. Another quantitative analysis is employing a nationally administered discrete choice experiment (DCE) survey to evaluate stakeholders’ preferences regarding provision of outpatient diagnostic services for staffle eye disease.

A separate secondary analysis will use the HERCULES patient-reported outcome measures from the exemplar clinic to generate a mapping algorithm to transform condition-specific (the Visual Function Index, VF-14) to generic (EQ-5D-5L) QOL outcomes, for use in interpreting the condition-specific measure more broadly.

#### Target Populations and eligibility

Recruitment targeted adult patients (over 18 years of age) attending follow-up clinic appointments within the Cataract, Glaucoma and Medical Retina services at existing Moorfields sites (City Road and Hoxton Eye-TRACs) and the new Moorfields at Brent Cross Eye-TRACs. Staff working within these clinics were also recruited for the quantitative and qualitative work.

For the Moorfields system-wide patient-level data for analysis of appointment delays, anonymised data from adult patients seen at Moorfields sites with staffle glaucoma or retinal disease were used.

To assess stakeholder preferences (in a discreet choice experiment) nationally/at national level, surveys were sent to adult ophthalmology patients in the UK (over 18 years of age), health care professionals and members of the general public.

### Data collection

#### Periods of recruitment: Follow up

The Eye-TRACs at Brent Cross operated initially for four 8-12 week iterations seeing approximately 600 patients per week, with spatial and operational reconfiguration between each cycle. In between iterations patients continued to attend the Eye-TRACs and clinics ran as normal.

Figure 1 demonstrates patients approached and declining participation in each iteration.

**Figure 1:**
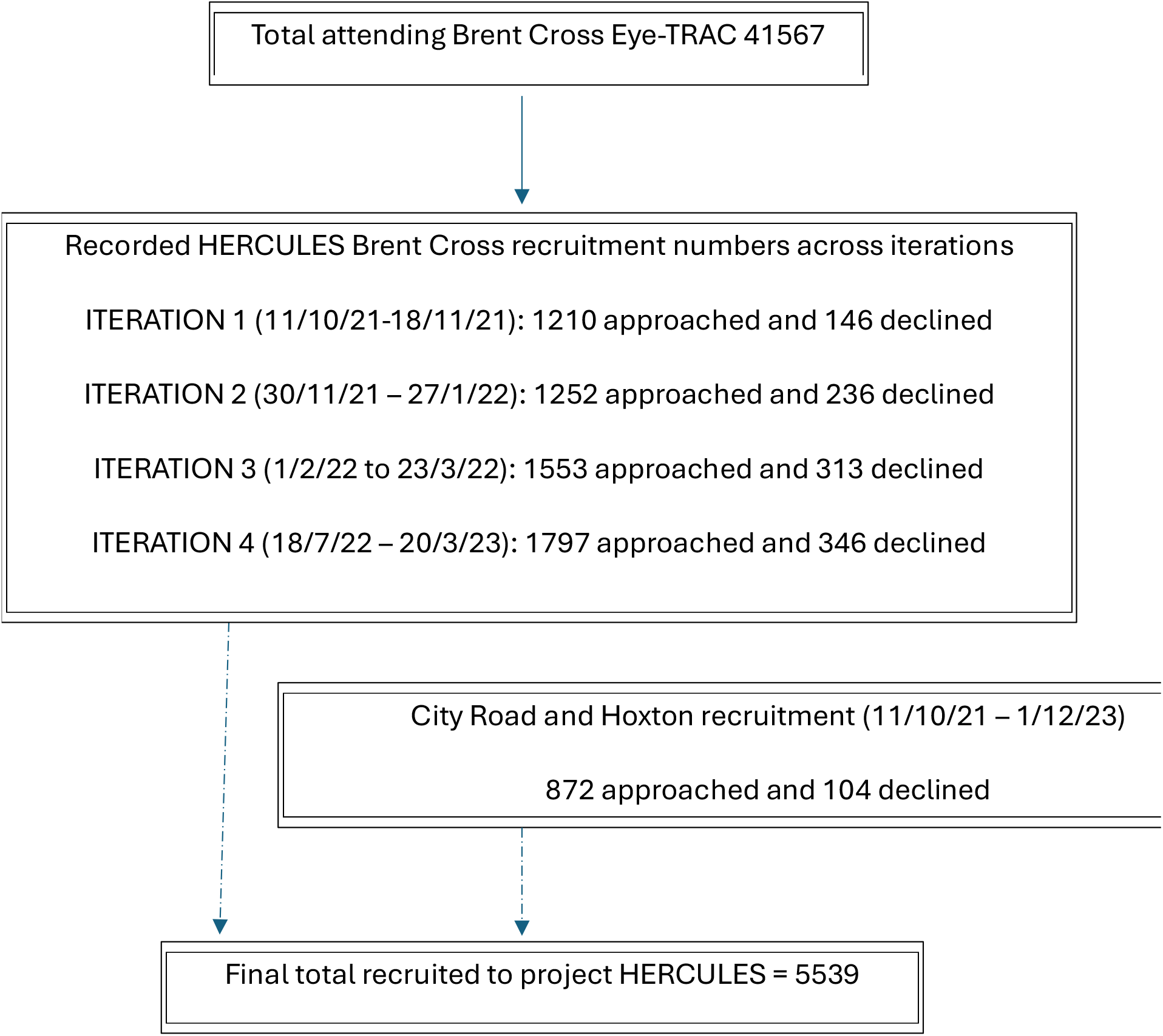
Flow chart showing numbers of patients approached by the recruitment team across iterations and numbers of patients declining participation.

Some documented reasons for non-participation were:

“not interested” – 297 patients

“no English / language barrier” – 144 patients “no time” – 27 patients

“negative emotions” (upset or angry on the day) – 5 patients.

#### Quantitative Data Collection and Analysis from Brent Cross

Participants attending Brent Cross who gave informed consent at the start of their clinic visit, were asked to complete the Visual Function Index (VF-14)(35) and the EQ-5D-5L(36, 37) with vision “bolt-on” as part of the PROMs questionnaire.

Patient positional monitoring during the spatial reconfigurations provided the following data:

1. Examination duration and patient journey times through the ultra-wideband real-time location tracking system (Ubisense).
2. Directly observed patient journey times with contextual information with subsequent analysis of spatial viewsheds in the clinics from every location to every other location based on principles of space syntax(38), highlighting patient experience, flows and overlaps or spatial bottlenecks.
3. Time stamps recorded by ophthalmic equipment, and patient arrival and departure times recorded by clerical staff.

These data will be analysed to triangulate the “centre of gravity” of patient journey times, relating these to the directly observed contextual information, and to perform mathematical modelling of operational processes and patient flow.

#### Qualitative Data Collection and Analysis

At the end of the scheduled clinic attendance, patients were asked to complete a brief service user questionnaire as part of the same PROMs questionnaire that evaluated patient experience within the facility and how this compared to any prior diagnostic clinics they may have attended.

The Brent Cross experimental clinic ran for a 26-month period and our dataset will permit comparisons of:

- Experiences of a cohort of patients who may have experienced two different configurations of Brent Cross, specifically:
  - Comparative experience of patients at both Brent Cross (research-focussed) and Hoxton (service-focussed) Eye-TRACs.
  - Experience of patients in both traditional “face to face” clinics and the Eye-TRACs.

The qualitative analysis (of participant interviews and surveys) delivered both rapid, formative learning and summative lessons on planning, delivery and experience of this innovation. Formative learning was provided via rapid qualitative analysis(39), operating in multiple cycles (reflecting the intervention being studied). Our data were drawn together using the Rapid Assessment Procedures (RAP) approach(40), which permits data collection and analysis to be conducted in parallel. The RAP sheets were updated after each instance of data collection (e.g. interview, meeting observation), facilitating quick and ongoing analysis and feedback with stakeholders.

A rapid ethnographic study to analyse unintended consequences (long appointments and missed diagnostic tests) employed semi-structured staff interviews, addressing the decision-making process in moving through the diagnostic pathway.

The summative analysis will be organised around two broad themes, reflecting our research questions addressing implementation, delivery and experience of these services. Our analyses combined inductive theory (theory building) and deductive (theory-guided) approaches.

1. Service delivery and experience: The first analysis will address service delivery and experiences of the Eye-TRACs studied, from staff and patient perspectives. Key themes will include a) enabling patient access to service (e.g. service location and pre-appointment communication), b) organisation and delivery of testing (e.g. influence of different spatial layouts and how patients are taken round the service) and c) managing difficulties in service delivery (e.g. technical and informational).
2. Planning and implementation: The second analysis will focus on the approaches to designing and implementing service innovations. Key themes will include a) how different clinical, managerial and academic perspectives combined to address design and implementation issues, b) how engagement with frontline staff influenced approaches through different iterations and c) how learning from earlier iterations shaped approaches in later iterations.

#### Data Management

We have and will continue to adhere to institutional information governance policies which comply with the UK General Data Protection Regulation (GDPR) and Data Protection Act (2018) at all times so that personal identifiable information (PII) is protected.

Retrospective electronic patient records from the wider Moorfields dataset of all glaucoma and medical retina patients who have been seen at any site of the Moorfields Eye Hospital (MEH) NHS Foundation Trust between 2018 and 2023, provided by Dun Jack Fu with assistance from members of the Moorfields Service Improvement and Sustainability team, have been anonymised and transferred securely into the UCL Data Safe Haven, and are being used for an interrupted time series analysis to assess the impact of opening the exemplar clinic on appointment delays. The Data Safe Haven is certified to the ISO27001 information security standard and conforms to NHS Digital’s Information Governance Toolkit.

Investigators at UCL were given data exports of the Brent Cross PROMs data containing only pseudonymised study data, following data quality checks by the Moorfields data management team. All study records will be retained for five years in secure storage within the Moorfields R&D offices following publication of the results.

#### Patient and Public Involvement and Engagement (PPIE)

PPIE was incorporated both in the iterative development of the Eye-TRACs and also in the design and implementation of the research methodology.

We have directly sought patients’ opinions for service development, throughout the iterative refinement of clinic design through qualitative surveys.

In terms of research and evaluation, one of the co-authors of this paper (SNa) is PPIE collaborator and was fully involved in the quantitative, qualitative and rapid ethnography sections of this project. SNa attends regular project meetings, contributes to development of research materials, discussions of progress in the work, interpretation of findings and is a co-author of research outputs.

The project as a whole is supported by three major patient-centred charities who share the vision of this project. These charities are Glaucoma UK, The Macular Society and Diabetes UK. The charities were involved in conception, design and funding of the project, and they will help with dissemination of findings to patient groups. They play a significant role in helping to develop the patient and public involvement as this iterative project progresses.

### FINDINGS TO DATE

41,567 patients attended the Moorfields Brent Cross Eye-TRAC between September 2021 and November 2023. Of these, 5,539 were recruited to Project HERCULES, including 2,199 medical retina patients, 2,993 glaucoma patients, and 347 cataract patients. Mean age of the cohort was 64.6 years (standard deviation 13.3 years,) minimum age was 17 years and maximum age was 98 years. Ethnicity profile reflects the diverse population of North London; 1444 patients were white British (26.1%,) 1,208 patients were Indian (22%,) 572 were black African or Caribbean (10.3%,) 1232 patients (22%) did not declare their ethnicity.

Table 1 summarises the attendances for each specialty during and in between the iterations. The numbers recruited for HERCULES for each specialty is included in italics with an asterisk*.

**TABLE 1:**
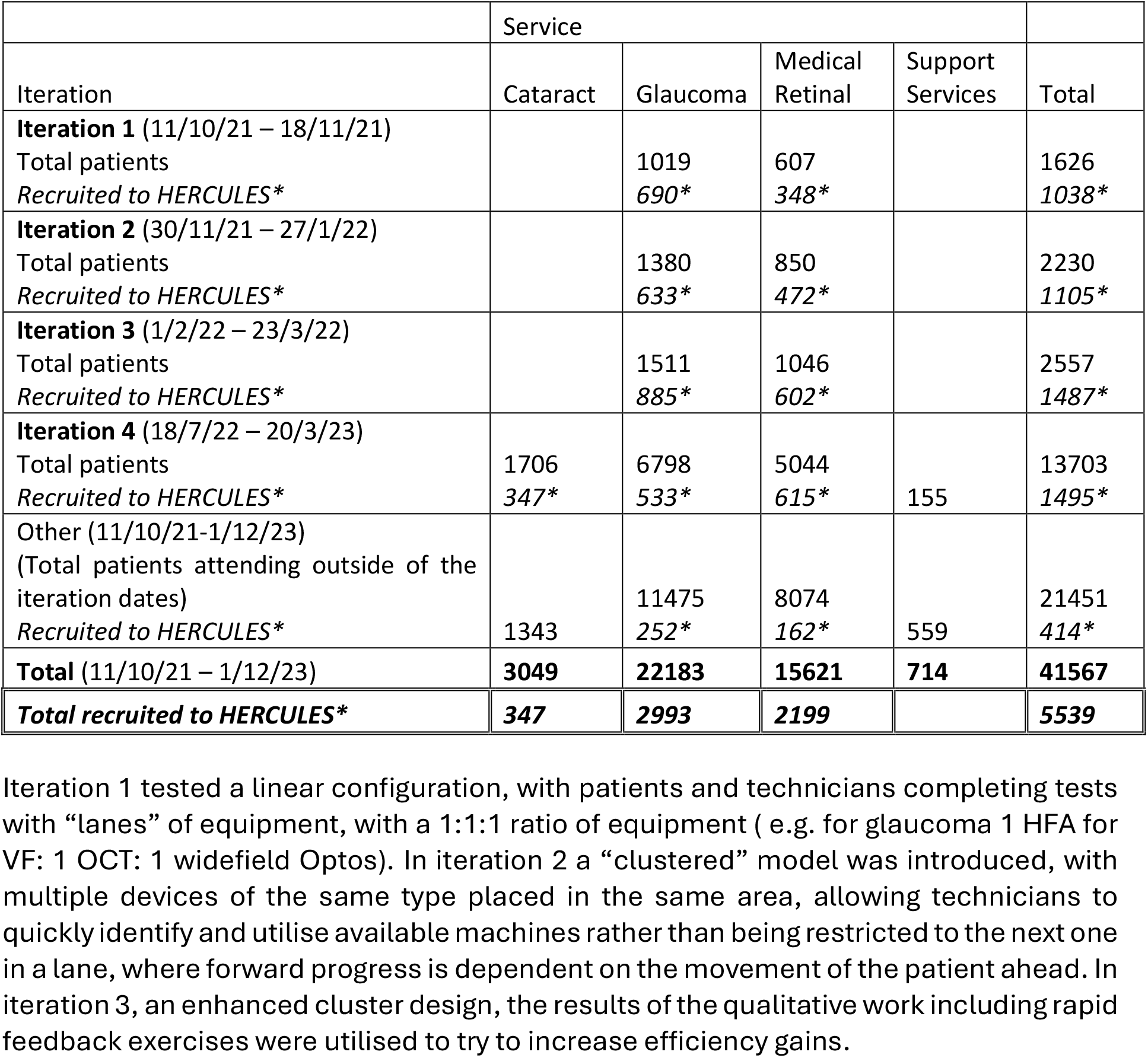
Patient numbers by service and date seen for NHS service at Moorfields Brent Cross.

Some of the rapid feedback exercises in iteration 4, examined the cause for excessively long appointments and found that complex patients who have significant additional care needs (either due to visual or other morbidity) were a major contributing factor to bottle-necks in the pathway. One interpretation is that to realise the potential efficiency gains of this method of working, dedicated diagnostic and monitoring clinics may be more suited, and more appropriately equipped, to deal with the larger number of mild/moderate disease severity cases, and that those with more complex disease, and systemic comorbidity, may be best cared for in a traditional hospital setting.

In iteration 4, “face to face” pre- and post-operative cataract clinics were introduced which created a more complex dynamic, and some disturbance in the smooth operating of the diagnostic and monitoring workstreams. We hypothesise that this was due to more complexity being introduced into the system and the loss of a consistent and predictable testing pathway that can occur when traditional face to face clinics are combined with Eye-TRACs. It demonstrates the need for understanding new models of care and the involvement of all stakeholders which emanates from research into healthcare planning and architecture through the development of new guidance.

### COLLABORATION

The findings of this research project are of significant relevance to NHS eye care and other high-volume ambulatory outpatient services, in the post COVID-19 era. Research findings will be shared with the Royal College of Ophthalmologists, regional Integrated Care Systems and the NHS Department for Outpatient Transformation to help inform planning for future diagnostic clinic models across the United Kingdom.

Our research output has been and will be submitted for presentation at national and international conferences and for peer-reviewed publication in the domains of ophthalmology, patient experience, health economics, healthcare, built environment design and human-computer interaction.

The findings will be shared with the wider patient population in collaboration with our partners, the Macular Society, Glaucoma UK and Diabetes UK who are the leading patient-focused charities relating to eye-care in the United Kingdom.

Duncan Wilson’s team have provided motion tracking data on this public URL: https://github.com/djdunc/hercules/tree/main/data/live.

All reasonable further data collaboration requests will be considered.

## FURTHER DETAILS

In February 2024, when the rental contract for the original unit expired, the Eye-TRACs at Brent Cross transitioned to a new service facility at a neighbouring unit, serving medical retina, glaucoma, cataract and keratoconus patients. The new clinic aims to deliver at least 600 appointments per week.

### Future work

Long term follow up of the HERCULES patient cohort is planned; to explore rates of treatment escalation and glaucoma progression in the patients seen in virtual clinics compared to face to face.

#### High Volume Low Complexity Cataract Services

Iteration 4 of Project HERCULES saw the introduction of “one-stop” clinician-patient “face-to-face” cataract clinics, to serve new and post-operative patients and list patients for surgery. Cataract surgery is the most performed elective surgical procedure in the UK and demand for this procedure is ever-increasing with an ageing population and particularly after disruption to elective surgical lists during the pandemic. In a bid to improve the overall efficiency and accessibility of healthcare services, high-volume low-complexity (HVLC) surgical hubs have been proposed as a potential solution to the backlog of patients waiting for elective surgeries. The HERCULES group seeks to define the optimal design parameters for high volume cataract surgical theatres in collaboration with researchers from the Bartlett School at UCL using lessons learned during HERCULES so far.

A further extension to the project includes a rapid realist review (RRR) to explore the influence of the HVLC cataract surgery services on patient, service, and system outcomes, and how these interplay with implementation strategies, workforce dynamics, and the patient experience.

#### IT refinement

Since the expansion of asynchronous “virtual” monitoring at Moorfields, we have found that clinical data review is one of the major challenges to efficiency in this care pathway. To address this, and to optimise processing efficiency and analysis of the large amount of clinical data and images acquired through the diagnostic clinic, researchers from the UCL Interaction Centre aim to create innovative strategies utilising artificial intelligence (AI) / machine learning (ML).

This team will explore innovative solutions based on large language model processing and utilise voice recognition software to integrate clinician commentary on virtual reviews into the structured electronic medical record, with a secondary focus on ensuring the new system can provide data that is easily useable in future health services research.

#### Generalisability and Application

We have demonstrated principles associated with higher numbers of patients examined in our Eye-TRACs serving patients with chronic, presumed staffle, eye disease, and believe these will be applicable to other high volume outpatient investigation and monitoring services beyond ophthalmology, both in the UK and globally. However, diberent cultural values in healthcare, and organisational environments, may create unforeseen operational variations or disruptions. Consequently, we are examining the impact of our clinic on waiting times across our NHS Trust’s network, and will work to build collaborations to further test our results and roll out this method of working across the UK and globally.

## Acknowledgements

We are grateful to Ms Jocelyn Cammack and Mrs Helen Baker, at NIHR Biomedical Research Centre at Moorfields Eye Hospital NHS Foundation Trust & UCL Institute of Ophthalmology, London, UK, for their guidance on public and patient involvement, and their participation in scientific discussions which shaped the project that we have outlined here. Ms Ella Preston, of Moorfields Service Improvement and Sustainability, Moorfields Eye Hospital NHS Foundation Trust, assisted with accessing patient activity data from hospital records. Paul Cartwright from the Moorfields Estates team and Jon Spencer the Chief Operating Officer of Moorfields provided operational support and logistical oversight across the iterations. Ubisense Ltd (Cambridge, UK) provided an ultrawideband tracking system at a discounted cost for use in this study. Optos Ltd (Dunfermline, UK) provided discounted wide field photography devices for use in this study. Zeiss provided OCT imaging devices and automated perimetry devices at a discounted cost for use in this study. Professor Sir Peng Khaw is supported by the Helen Hamlyn Trust, the Nolan Family, the Katz Foundation, Moorfields Eye Charity and the NIHR Biomedical Research Centre at Moorfields Eye Hospital and the UCL Institute of Ophthalmology. The views expressed in this paper are those of the authors and not necessarily those of any funding body or the Department of Health.

## Collaborators

### HERCULES Consortium: (FULL LIST)

Aadil Kazi – NIHR BRC, Angus Ramsay – UCL, Anne Symons – UCL, Connor Beddow - Moorfields NHS FT, Chris Leak - Moorfields NHS FT, Caroline S Clarke – UCL, Dun Jack Fu - Moorfields NHS FT, Duncan Wilson – UCL, Dhakshi Muhundhakumar - Moorfields NHS FT, Lina Song – UCL, Declan Flanagan - Moorfields NHS FT, Elisha Chung - Moorfields NHS FT, Ella Preston - Moorfields NHS FT, Farbod Afshar Bakeshloo – UCL, Giovanni Ometto – UCL, Gus Gazzard - Moorfields NHS FT, Grant Mills - UCL, George Damianidis - Moorfields NHS FT, Helen Baker - Moorfields NHS FT, Hari Jayaram - Moorfields NHS FT, Ian Eames – UCL, Iqbal Fahmi – UCL, Irinie Roufaeel – UCL, Jocelyn Cammack - Moorfields NHS FT, Josefine Magnusson - UCL, Jemima Unwin – UCL, Jonathan Wilson - Moorfields NHS FT, Joy Adesanya - Moorfields NHS FT, Kerstin Sailer – UCL, Kathryn Scotcher - Moorfields NHS FT, Kimberley Quan - Moorfields NHS FT, Martin Utley – UCL, Matala Dyke - Moorfields NHS FT, Naomi J Fulop – UCL, Natalie O’Shea - Moorfields NHS FT, Paul Foster – UCL, Peter Scully - UCL, Paul Webster – Ubisense, Paula Lorgelly – University of Auckland, Pei Li Ng – UCL, Peter Thomas - Moorfields NHS FT, Rachel Thompson - Moorfields NHS FT, Robin Hamilton - Moorfields NHS FT, Rosica Pachilova – UCL, Rouba Ibrahim – UCL, Susana Frazao Pinheiro – UCL, Siyabonga Ndwandwe – UCL, Saheli Gandhi – UCL, Samiul Alom - Moorfields NHS FT, Sherene Ettiene - Moorfields NHS FT, Sobha Sivaprasad - Moorfields NHS FT, Stacey Angus - Moorfields NHS FT, Steve Napier - Moorfields NHS FT, Yue Tang – UCL, Dominika - UCL, Ecem – UCL, Xiaoming Li– UCL, Muna Ayah - Moorfields NHS FT, Nadine Abdelgalil - Moorfields NHS FT, Paul Cartwright - Moorfields NHS FT, Sarah Davies - Moorfields NHS FT, Sandi Drewett - Moorfields NHS FT, Clare Feasby - Moorfields NHS FT, Simranjit Gill - Moorfields NHS FT, Steven Gill - Moorfields NHS FT, Nick Hardie - Moorfields NHS FT, Jamie Henderson - Moorfields NHS FT, Lesley Henry - Moorfields NHS FT, Peng Tee Khaw - Moorfields NHS FT, Richard Lee - Moorfields NHS FT, Sarah Martin - Moorfields NHS FT, Mary Masih - Moorfields NHS FT, Luke Nicholson - Moorfields NHS FT, Tulga Reis - Moorfields NHS FT, Nick Roberts - Moorfields NHS FT, Ana Sanchez - Moorfields NHS FT, Jon Spencer - Moorfields NHS FT, Karen Titmus - Moorfields NHS FT, Eleanor Dean – Akeso, Nick Hynes – SOMO Global, Olivia Jeffrey - Akeso, Chris Robson - Akeso, Tom Blair – Ubisense, Nick Burt – UCL, Dolores Conroy – UCL

## Contributors

Critical revision of the manuscript for important intellectual content: DM, PF, HJ, SS, CSC, AIGR, GM, AS, KS. PJF, DM, HJ, SS, SNd, CSC, AR, GM had access to the relevant sections of the data in the study. Concept and design: PJF, HJ, SS, CSC, AIGR, SNa, GM, AS, PS, DW. Data acquisition and analysis: HERCULES clinic data GO, DM, PF, DJF. Ubisense data obtained by DW and team, DOTS data obtained by RP, KS. Obtained funding: PJF, HJ, SS, PTK, JW. All authors approved the final manuscript.

## Patient and public involvement

Patients and/or the public were involved in the design, conduct and dissemination plans of this research. Refer to the Methods section for further details.

## Patient consent for publication

Not applicable

## Ethics approval

Project HERCULES was approved by North East - York NHS Research Ethics Committee (IRAS 303760)

## Data availability statement

Professor Duncan Wilson’s team have provided motion tracking data on this public URL: https://github.com/djdunc/hercules/tree/main/data/live All reasonable further data collaboration requests will be considered.

